# Pre-Existing Inflammatory Disease Predicts Cutaneous Immunotherapy Toxicity Development: A Multi-Institutional Cohort Study

**DOI:** 10.1101/2023.08.28.23294695

**Authors:** Guihong Wan, Nga Nguyen, Bonnie W. Leung, Hannah Rashdan, Kimberly Tang, Katie Roster, Michael R. Collier, Pearl O. Ugwu-Dike, Neel S. Raval, Nora A. Alexander, Ruple Jairath, Jordan Phillipps, Munachimso Amadife, Shijia Zhang, Alexander Gusev, Steven T. Chen, Kerry L. Reynolds, Nicole R. LeBoeuf, Shawn G. Kwatra, Yevgeniy R. Semenov

**Affiliations:** Massachusetts General Hospital, Department of Dermatology, Boston, MA; Harvard Medical School, Department of Dermatology, Boston, MA; Dana Farber Cancer Institute, Department of Medicine, Boston, MA; Massachusetts General Hospital, Department of Medicine, Division of Oncology, Boston, MA; Dana Farber Cancer Institute, Department of Dermatology, Boston, MA; Johns Hopkins University, Department of Dermatology, Baltimore, MD

**Author notes:** **Corresponding author:** Yevgeniy R. Semenov, MD, MA, Department of Dermatology, Massachusetts General Hospital, Harvard Medical School, 40 Blossom Street, Bartlett Hall 6R, Room 626, Boston, MA 02114. Designate co-first authors (Guihong Wan, Nga Nguyen). Designates co-senior authors (Shawn G. Kwatra, Yevgeniy R. Semenov). Twitter handle: @EugeneSemenovMD. **Reprint requests:** Yevgeniy R. Semenov, MD, MA. **IRB approval status:** Reviewed and approved by Mass General Brigham Institutional Review Board (Protocol # 2020P002307). **Patient consent on file:** N/A.

**Keywords:** pre-existing inflammatory disease, autoimmunity, cutaneous immune-related adverse events, cutaneous adverse reactions, immunotherapy, immune checkpoint inhibitor, oncodermatology

## Abstract

**Background:** Relationships between pre-existing inflammatory diseases (pIDs) and cutaneous immune-related adverse events (cirAEs) have not been well-studied. This study is to investigate associations between pIDs and cirAEs among immune-checkpoint inhibitor (ICI) recipients at the Mass General Brigham healthcare system.

**Methods:** Electronic health records were reviewed to ascertain cirAE status. Patients’ pID status was determined using International Classification of Diseases (ICD) codes. Cox proportional hazard, logistic regression, and linear regression models were performed.

**Results:** Among 3607 ICI recipients, 1354 had pIDs, and 672 developed cirAEs. After covariate adjustments, patients with cutaneous pIDs (HR:1.56, p<0.001) or both cutaneous and non-cutaneous pIDs (HR:1.76, p<0.001) had increased cirAE risk in contrast to patients with non-cutaneous pIDs alone (HR:1.01, p=0.9). In adjusted ordinal logistic regression modeling, cutaneous pIDs (OR:1.55, p<0.0001) and the presence of both cutaneous pIDs and non-cutaneous pIDs (OR:1.71, p=0.002) were associated with increased cirAE severity. The time to cirAE onset was different between the cutaneous pID group and the non-cutaneous pID group (Mean: 98 vs. 146 days, p=0.021; Beta: -0.11, p=0.033).

**Conclusions:** ICI recipients with cutaneous pIDs should have increased clinical monitoring due to their increased risk of cirAE development, severity, and earlier onset.

---

Though immune checkpoint inhibitors (ICIs) have considerably improved patient outcomes, they are associated with morbid and potentially life-threatening immune-related adverse events (irAEs). As a result of the immune-mediated nature of these toxicities and despite recent evidence that ICI recipients with pre-existing inflammatory diseases (pIDs) are not at increased risk of mortality ^2,3^, patients with pIDs have been largely excluded from ICI clinical trials, and data on toxicity development in this population is lacking.^1^ To address this question, we leverage a large multi-institutional cohort of ICI recipients to examine associations of pIDs with cutaneous irAEs (cirAEs), which are the most common and earliest toxicities to occur with ICI therapy.

The presence, severity, and timing of cirAEs were ascertained by manual chart review using previously published approaches (**eMethod 1**).^4,5^ Patients’ pID status was determined using International Classification of Diseases (ICD) codes according to consensus definitions for inflammatory disorders (**eMethod 2)**. Multivariate Cox Proportional Hazard (CoxPH) models were utilized to investigate relationships between pIDs and cirAEs. Hazard Ratios (HRs) and 95% confidence intervals (CIs) were computed. Adjusted logistic and linear regressions were used to examine associations of pIDs with cirAE severity and timing.

We identified 3,607 ICI recipients at the Massachusetts General Brigham (MGB) (**eFigure 1)**. Among them, 1,354 (37.5%) had pIDs, and 672 (18.6%) developed cirAEs (**eTable 3)**. In the pID group, 517 (38.2%) had one or more cutaneous pIDs (c-pIDs), and 837 (61.8%) had only non-cutaneous pIDs (nc-pIDs). Multivariate CoxPH modeling suggests that patients with pIDs were more likely to develop cirAEs (HR:1.20, 95% CI: [1.01, 1.42], p=0.036) (**Table 1**) compared to patients without pIDs. Specifically, c-pIDs increased the cirAE risk (HR:1.56, 95% CI: [1.25, 1.94], p<0.001), whereas nc-pIDs did not (HR:1.01, 95% CI: [0.82, 1.25], p=0.9). The cirAE risk was highest for patients with both c-pIDs and nc-pIDs (HR:1.76, 95% CI: [1.28, 2.42], p<0.001). This was most significant among patients with specific pre-existing dermatologic conditions, including atopic dermatitis (HR:3.58, 95% CI: [2.34, 5.48], p<0.001), morphea (HR: 3.23, 95% CI: [1.42, 7.37], p=0.005), and psoriasis (HR:1.79, 95% CI: [1.23, 2.62], p=0.003). Sensitivity analyses yielded similar conclusions **(eTables 5-6)**.

**Table 1.**
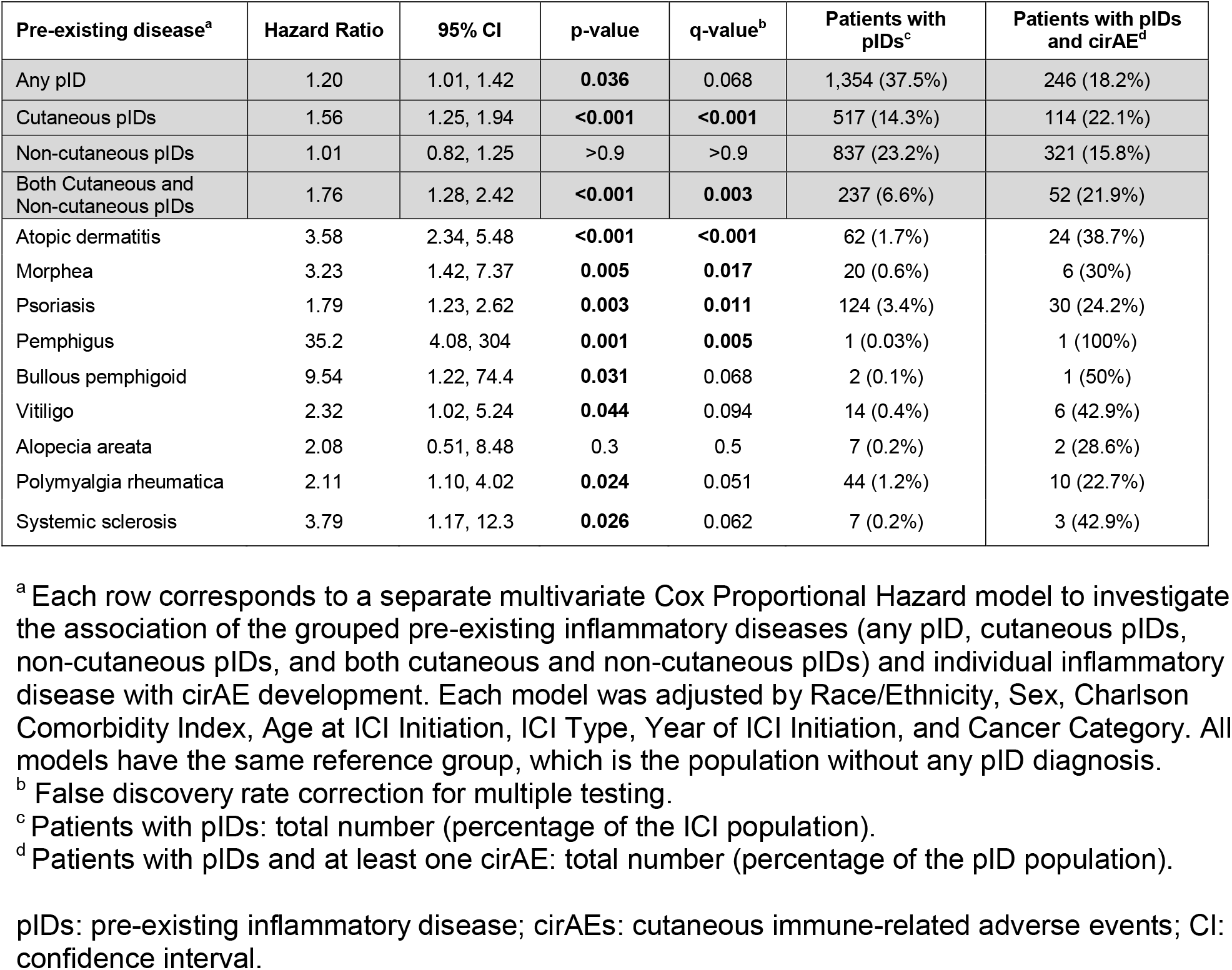
Associations of pIDs with cirAE development.

| Pre-existing disease<sup>a</sup> | Hazard Ratio | 95% CI | p-value | q-value<sup>b</sup> | Patients with pIDs<sup>c</sup> | Patients with pIDs and cirAE<sup>d</sup> |
| --- | --- | --- | --- | --- | --- | --- |
| Any pID | 1.20 | 1.01, 1.42 | **0.036** | 0.068 | 1,354 (37.5%) | 246 (18.2%) |
| Cutaneous pIDs | 1.56 | 1.25, 1.94 | **<0.001** | **<0.001** | 517 (14.3%) | 114 (22.1%) |
| Non-cutaneous pIDs | 1.01 | 0.82, 1.25 | >0.9 | >0.9 | 837 (23.2%) | 321 (15.8%) |
| Both Cutaneous and Non-cutaneous pIDs | 1.76 | 1.28, 2.42 | **<0.001** | **0.003** | 237 (6.6%) | 52 (21.9%) |
| Atopic dermatitis | 3.58 | 2.34, 5.48 | **<0.001** | **<0.001** | 62 (1.7%) | 24 (38.7%) |
| Morphea | 3.23 | 1.42, 7.37 | **0.005** | **0.017** | 20 (0.6%) | 6 (30%) |
| Psoriasis | 1.79 | 1.23, 2.62 | **0.003** | **0.011** | 124 (3.4%) | 30 (24.2%) |
| Pemphigus | 35.2 | 4.08, 304 | **0.001** | **0.005** | 1 (0.03%) | 1 (100%) |
| Bullous pemphigoid | 9.54 | 1.22, 74.4 | **0.031** | 0.068 | 2 (0.1%) | 1 (50%) |
| Vitiligo | 2.32 | 1.02, 5.24 | **0.044** | 0.094 | 14 (0.4%) | 6 (42.9%) |
| Alopecia areata | 2.08 | 0.51, 8.48 | 0.3 | 0.5 | 7 (0.2%) | 2 (28.6%) |
| Polymyalgia rheumatica | 2.11 | 1.10, 4.02 | **0.024** | 0.051 | 44 (1.2%) | 10 (22.7%) |
| Systemic sclerosis | 3.79 | 1.17, 12.3 | **0.026** | 0.062 | 7 (0.2%) | 3 (42.9%) |

<sup>a</sup> Each row corresponds to a separate multivariate Cox Proportional Hazard model to investigate the association of the grouped pre-existing inflammatory diseases (any pID, cutaneous pIDs, non-cutaneous pIDs, and both cutaneous and non-cutaneous pIDs) and individual inflammatory disease with cirAE development. Each model was adjusted by Race/Ethnicity, Sex, Charlson Comorbidity Index, Age at ICI Initiation, ICI Type, Year of ICI Initiation, and Cancer Category. All models have the same reference group, which is the population without any pID diagnosis.
<sup>b</sup> False discovery rate correction for multiple testing.
<sup>c</sup> Patients with pIDs: total number (percentage of the ICI population).
<sup>d</sup> Patients with pIDs and at least one cirAE: total number (percentage of the pID population).
pIDs: pre-existing inflammatory disease; cirAEs: cutaneous immune-related adverse events; CI: confidence interval.

Furthermore, c-pIDs were associated with a higher risk of severe cirAEs (**Table 2**). In adjusted ordinal logistic regression modeling, c-pIDs (OR:1.55, 95% CI: [1.22, 1.96], p<0.0001) and the presence of both c-pIDs and nc-pIDs (OR:1.71, 95% CI: [1.21, 2.38], P=0.002) were associated with increased cirAE severity. The time to cirAE onset was different between the c-pID group and the nc-pID group (Mean: 98 vs. 146 days, p=0.021; Beta: -0.11, 95% CI: [-0.21, -0.01], p=0.033) (**eTable 4**).

**Table 2.** Impact of pIDs on the cirAE severity using logistic regression models.

|  | **CirAE Severity Grade<sup>a</sup>** | **Odds Ratio** | **95% CI** | **p-value** |
| --- | --- | --- | --- | --- |
| Cutaneous pIDs | >=3 | 1.72 | 1.04, 2.76 | 0.029 |
|  | >=2 | 1.60 | 1.18, 2.15 | 0.002 |
|  | >=1 | 1.57 | 1.23, 2.00 | <0.001 |
|  | Overall ordinal regression<sup>b</sup> | 1.55 | 1.22, 1.96 | <0.001 |
| Both Cutaneous and Non-cutaneous pIDs | >=3 | 2.34 | 1.21, 4.25 | 0.008 |
|  | >=2 | 1.90 | 1.25, 2.84 | 0.002 |
|  | >=1 | 1.69 | 1.19, 2.37 | 0.003 |
|  | Overall ordinal regression<sup>b</sup> | 1.71 | 1.21, 2.38 | 0.002 |

<sup>a</sup> Each row corresponds to a separate binomial or ordinal logistic regression model. Each model was adjusted by Race/Ethnicity, Sex, Charlson Comorbidity Index, Age at ICI Initiation, ICI Type, Year of ICI Initiation, and Cancer Category. All binomial models (>=3, >=2, and >=1) have the same reference group, which is the population without any pID diagnosis.
<sup>b</sup> Scale of cirAE severity: 1-4; No cirAE: 0.
pIDs: pre-existing inflammatory disease; cirAE: cutaneous immune-related adverse event; CI: confidence interval.

This study demonstrated that increased cirAE risk was observed specifically in patients with c-pIDs, suggesting that immune-mediated predisposition to cirAE development is organ specific with possible tissue-specific or HLA-associated immunogenetic susceptibilities. Moreover, we identified an accentuated risk for high-grade (grade 3 or higher) and earlier onset cirAEs among patients with c-pIDs compared to those with nc-pIDs. The limitations of our study include use of ICD codes for pID status and retrospective grading of cutaneous eruptions.

## Supporting information

Supplemental Tabels

## Data Availability

All relevant data are available from the corresponding author. All summary data supporting the findings of this study are available within the article and/or its supplementary materials. The patient data generated for this study can only be shared per specific institutional review board (IRB) requirements. Upon a request to the corresponding author, a data-sharing agreement can be initiated following institution-specific guidelines.

