## Supplemental Tabels for "Pre-Existing Inflammatory Disease Predicts Cutaneous Immunotherapy Toxicity Development: A Multi-Institutional Cohort Study"

### Supplementary Material

**Total number of eMethods: 2**
**eMethod 1.** Assessment of cirAE status.
**eMethod 2.** Determination of pID status.

**Total number of eFigures: 1**
**eFigure 1.** Data collection flow diagram.

**Total number of eTables: 6**
**eTable 1.** ICD-9 and ICD-10 codes used to identify cutaneous pIDs.
**eTable 2.** ICD-9 and ICD-10 codes used to identify non-cutaneous pIDs.
**eTable 3.** Characteristics of the study population.
**eTable 4.** Impact of pIDs on cirAE timing.
**eTable 5.** Associations of pIDs with cirAE development using multivariate Cox Proportional Hazards models, where at least two instances of each pID ICD code were required to be categorized as a pID.
**eTable 6.** Associations of pIDs with cirAE development using multivariate Cox Proportional Hazards models, where only the pID ICD codes within 6 months or 12 months before ICI initiation were included.

**eMethod 1.** Assessment of cirAE status.

| Likelihood | Clinical features | CirAE |
|---|---|---|
| 1<br>(highly unlikely) | Flare of preexisting conditions, or if cutaneous eruption is more likely to be an infusion reaction | No |
| 2<br>(unlikely) | Cutaneous eruption more likely attributed to a concurrent medication | No |
| 3<br>(likely) | Cutaneous eruption more likely attributed to immune checkpoint inhibitor | Yes |
| 4<br>(highly likely) | Cutaneous eruption more likely attributed to immune checkpoint inhibitor with supportive evidence by dermatology evaluation, skin biopsy, and/or photographs | Yes |

We defined our patient population by identifying patients who received ICI therapy from billing records. Manual chart reviews were conducted by two independent reviewers to ascertain the presence, severity grade, and timing of cirAEs following ICI therapy initiation. A third reviewer arbitrated the instances where the two reviewers did not achieve concordance.

CirAE phenotyping was completed in accordance with the definitions for dermatologic immune-related adverse events in a recent study.[3] A score of 1 (highly unlikely) to 4 (highly likely) of cirAEs was assigned to each patient. The score was determined by a manual review of patient charts and documentation, including images, morphology, histology, timing, competing medications, course of the rash, and response to treatment. Patients with likelihood scores of 3 and 4 were categorized as having a cirAE.

- A highly likely score (4) was assigned if there was clinician documentation for immune-checkpoint inhibitor-related etiology, appropriate timing, and morphology as defined in the above modified-Delphi consensus statement on cirAE definitions and prior literature on cirAEs, no competing medications that would contribute to the development of cutaneous eruptions, and/or supportive evidence via photographs, histology, dermatologist or oncodermatologist evaluation.
- A likely score (3) was assigned if there were competing medications present, but the competing medication is not likely to be the trigger given the timing or morphology of cirAE in relation to the competing medication.
- An unlikely score (2) was assigned if, given the timing of the ICI, there was a competing trigger more likely to have caused the eruption in comparison to ICI.
- A highly unlikely score (1) was assigned if the timing of the cutaneous eruption did not coincide with the timing of ICI or if another cause for the cutaneous eruption was favored by the dermatologist.

Suspected events were graded on a scale of 1 (mild) to 4 (life-threatening) using Common Terminology Criteria for Adverse Events version 5.0.[4]

**eMethod 2.** Determination of pID status.

The ninth and tenth versions of International Classification of Diseases (ICD) codes were used to determine a patient's pID status:

- We collected a list of inflammatory conditions as specified in **eTable 1** and **eTable 2**. They were extracted from a comprehensive list of autoimmune diseases: https://autoimmune.org/disease-information, and refined based on literature and expert consensus [NRL, SGK, YRS].
- If a patient had any pre-existing (or any history of) inflammatory disease diagnosis before ICI initiation (**eTable 1** and **eTable 2**), then this patient is labeled as having pIDs; otherwise, this patient is labeled as having no pIDs.
- We further characterized the pID type. A patient's pID category is "cutaneous pID" if the patient was diagnosed with at least one pre-existing cutaneous conditions included in **eTable 1**. The pID category is "non-cutaneous pIDs" if the patient was diagnosed with at least one pre-existing disease included in **eTable 2** but not any pre-existing cutaneous disease included in **eTable 1**.
- We also considered patients diagnosed with at least one disease included in **eTable 1** and at least one disease in **eTable 2** (both cutaneous and non-cutaneous pID group).

Sensitivity analyses were performed to evaluate the robustness of the results to the definition of pIDs. The primary cohort required at least one instance of each pID ICD code to be categorized as a pID. In the sensitivity cohort, at least two instances of each pID ICD code were required to be categorized as a pID. We also conducted experiments where only the pID ICD codes within 6 months or 12 months before ICI initiation were included.

**eTable 1.** ICD-9 and ICD-10 codes used to identify cutaneous pIDs.

| Index | Disease | ICD-10 | ICD-9 |
|---|---|---|---|
| 1 | Dermatomyositis | M33 (Dermatopolymyositis) | 710.3 (Dermatomyositis) |
| 2 | Psoriasis | L40 (psoriasis) | 696.0 (arthropathic psoriasis)<br>696.1 (Other psoriasis) |
| 3 | Lichen planus | L43 (Lichen planus) | 697.0 (Lichen planus) |
| 4 | Vitiligo | L80 (Vitiligo) | 709.01 (Vitiligo) |
| 5 | Bullous pemphigoid | L12 (Pemphigoid) | 694.5 (Pemphigoid) |
| 6 | Pemphigus | L10 (Pemphigus) | 694.4 (Pemphigus) |
| 7 | Atopic dermatitis | L20 (Atopic dermatitis) | 691.8 (Other atopic dermatitis and related conditions) |
| 8 | Seborrheic dermatitis | L21 (Seborrheic dermatitis) | 690.1 (Seborrheic dermatitis) |
| 9 | Lupus erythematosus | L93 (Lupus erythematosus) | 695.4 (Lupus erythematosus) |
| 10 | Alopecia areata | L63 (Alopecia areata) | 704.09 (Other alopecia) |
| 11 | Morphea | L90.0 (Lichen sclerosus et atrophicus)<br>L94.0 (Localized scleroderma)<br>L94.3 (Sclerodactyly) | 701.0 (Circumscribed scleroderma) |
| 12 | Dermatitis herpetiformis | L13.0 (Dermatitis herpetiformis) | 694.0 (Dermatitis herpetiformis) |
| 13 | Skin vasculitis | L95.8 (Other vasculitis limited to the skin)<br>L95.9 (Vasculitis limited to the skin, unspecified) | 709.1 (Vascular disorders of skin) |
| 14 | Mucositis | K12 (Stomatitis and related lesions) | 528.00 (Stomatitis and mucositis, unspecified)<br>528.09 (Other stomatitis and mucositis)<br>528.2 (Oral aphthae) |
| 15 | Pyoderma gangrenosum | L88 (Pyoderma gangrenosum) | 686.01 (Pyoderma gangrenosum) |
| 16 | Hidradenitis suppurativa | L73.2 (Hidradenitis suppurativa) | 705.83 (Hidradenitis) |
| 17 | Erythema nodosum | L52 (Erythema nodosum) | 695.2 (Erythema nodosum) |
| 18 | Autoimmune urticaria | L50.8 (Other urticaria) | 708.8 (Other specified urticaria) |
| 19 | Linear IgA | L13.8 (Other specified bullous disorders) | 694.8 (Other specified bullous dermatoses) |

**eTable 2.** ICD-9 and ICD-10 codes used to identify non-cutaneous pIDs.

| Index | Disease | ICD-10 | ICD-9 |
|---|---|---|---|
| 1 | Rheumatoid arthritis | M05 (Rheumatoid arthritis with rheumatoid factor)<br>M06 (Other rheumatoid arthritis)<br>M08 (Juvenile rheumatoid arthritis) | 714.0 (Rheumatoid arthritis) |
| 2 | Systemic sclerosis | M34 (Systemic sclerosis) | 710.1 (Systemic sclerosis) |
| 3 | Systemic Lupus | M32 (Systemic lupus erythematosus) | 710.0 (Systemic lupus erythematosus) |
| 4 | Inflammatory bowel disease | K50 (Crohn's disease)<br>K51 (Ulcerative colitis)<br>K52 (Other and unspecified noninfective gastroenteritis and colitis) | 555.9 (Crohn's disease of unspecified site)<br>556.6 (Universal ulcerative chronic colitis)<br>556.9 (Ulcerative colitis, unspecified)<br>558.9 (Other and unspecified noninfectious gastroenteritis and colitis) |
| 5 | Ankylosing spondylitis | M45 (Ankylosing Spondylitis) | 720.0 (Ankylosing spondylitis) |
| 6 | Sicca syndrome | M35.0 (Sjögren syndrome) | 710.2 (Sicca syndrome) |
| 7 | Polymyalgia rheumatica | M35.3 (Polymyalgia rheumatica) | 725 (Polymyalgia rheumatica) |
| 8 | Mixed connective tissue | M35.9 (Systemic involvement of connective tissue, unspecified) | 710.9 (Unspecified diffuse connective tissue disease) |
| 9 | Type 1 diabetes | E10 (Type 1 diabetes mellitus) | 250.x1 (Diabetes mellitus, type I, not stated as uncontrolled)<br>250.x3 (Diabetes mellitus, type I, uncontrolled) x = 0 to 9. |
| 10 | Myasthenia gravis | G70.0 (Myasthenia gravis) | 358.00 (Myasthenia gravis without acute exacerbation)<br>358.01 (Myasthenia gravis with acute exacerbation) |
| 11 | Graves' disease | E05.00 (Thyrotoxicosis with diffuse goiter without thyrotoxic crisis or storm)<br>E05.01 (Thyrotoxicosis with diffuse goiter with thyrotoxic crisis or storm) | 242.00 (Toxic diffuse goiter with no crisis or storm)<br>242.01 (Toxic diffuse goiter crisis or storm) |
| 12 | Autoimmune thyroiditis | E06.3 (Autoimmune thyroiditis) | 245.2 (Chronic lymphocytic thyroiditis) |
| 13 | Addison’s disease | E27.1 (Addison’s disease)<br>E27.2 (Addisonian crisis)<br>E27.3 (Drug-induced adrenocortical insufficiency)<br>E27.4 (Other and unspecified adrenocortical insufficiency) | 255.41 (Glucocorticoid deficiency) |
| 14 | Autoimmune Hepatitis | K75.4 (Autoimmune hepatitis) | 571.42 (Autoimmune hepatitis) |
| 15 | Celiac disease | K90.0 (Celiac disease) | 579.0 (Celiac disease) |

| | | | |
|---|---|---|---|
| 16 | Vasculitis | I77.6 (Arteritis, unspecified)<br>M30.1 (Polyarteritis with lung involvement)<br>M31 (Other necrotizing vasculopathies) | 446.0 (Polyarteritis nodosa)<br>446.20 (Hypersensitivity angiitis, unspecified)<br>446.21 (Goodpasture's syndrome)<br>446.29 (Other specified hypersensitivity angiitis)<br>446.3 (Lethal midline granuloma)<br>446.4 (Wegener's granulomatosis)<br>446.5 (Giant cell arteritis)<br>446.7 (Takayasu's disease)<br>447.5 (Necrosis of artery)<br>447.6 (Arteritis, unspecified) |
| 17 | Sarcoidosis | D86 (Sarcoidosis) | 135 (Sarcoidosis) |
| 18 | Autoimmune hemolytic anemia | D59.0 (Drug-induced autoimmune hemolytic anemia)<br>D59.1 (Other autoimmune hemolytic anemias) | 283.0 (Autoimmune hemolytic anemias) |
| 19 | Biliary cirrhosis | K74.3 (Primary biliary cirrhosis)<br>K74.4 (Secondary biliary cirrhosis)<br>K74.5 (Biliary cirrhosis, unspecified) | 571.6 (Biliary cirrhosis) |
| 20 | Guillain-barre syndrome | G61.0 (Guillain-barre syndrome) | 357.0 (Guillain-barre syndrome) |
| 21 | Reiter’s disease | M02.3 (Reiter’s disease) | 099.3 (Reiter’s disease) |
| 22 | Hypothyroidism | E03.8 (Other specified hypothyroidism)<br>E03.9 (Hypothyroidism, unspecified) | 244.8 (Other specified acquired hypothyroidism)<br>244.9 (Hypothyroidism, unspecified) |
| 23 | Multiple Sclerosis | G35 (Multiple Sclerosis) | 340 (Multiple Sclerosis) |

**eFigure 1.** Data collection flow diagram

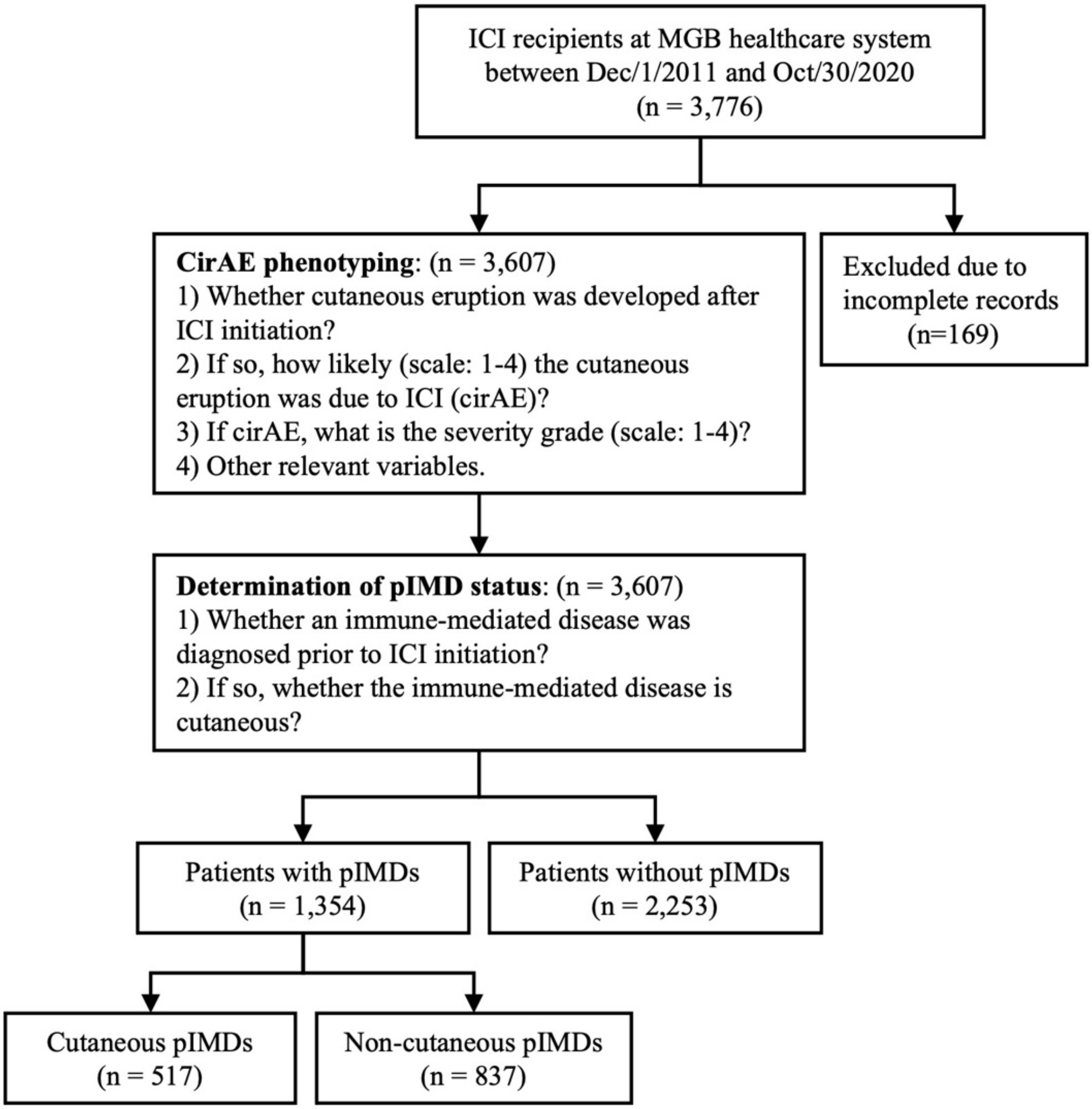

**eTable 3**. Characteristics of the study population.

| Characteristic | All patients (N=3,607) | pID Condition | | |
|---|---|---|---|---|
| | | pID N=1,354 (37.5%) | No pID N=2,253 (62.5%) | P-value |
| **CirAE Status** | | | | |
| No CirAE | 2935 (81.4%) | 1108 (81.8%) | 1827 (81.1%) | 0.611 |
| CirAE | 672 (18.6%) | 246 (18.2%) | 426 (18.9%) | |
| **Time to CirAE[a] (days)** | | | | |
| Mean (SD) | 131 (176) | 124 (169) | 134 (180) | 0.458 |
| Median [IQR] | 64 [21, 175] | 63 [21, 154] | 66 [21, 182] | |
| **CirAE Severity Grade** | | | | |
| 1 | 289 (8.0%) | 101 (7.5%) | 188 (8.3%) | 0.651 |
| 2 | 254 (7.0%) | 100 (7.4%) | 154 (6.8%) | |
| >=3[b] | 129 (3.6%) | 45 (3.3%) | 84 (3.7%) | |
| **pID Category** | | | | |
| Cutaneous pIDs | 517 (14.3%) | 517 (38.2%) | N/A | |
| Non-cutaneous pIDs | 837 (23.2 %) | 837 (61.8%) | N/A | |
| No pIDs | 2253 (62.5%) | N/A | 2253 (100%) | |
| **Sex** | | | | |
| Female | 1637 (43.4%) | 681 (50.3%) | 956 (42.4%) | <0.001 |
| Male | 1970 (54.6%) | 673 (49.7%) | 1297 (57.6%) | |
| **Race/Ethnicity** | | | | |
| White | 3274 (90.8%) | 1231 (90.9%) | 2043 (90.7%) | 0.060 |
| Asian | 130 (3.6%) | 44 (3.2%) | 86 (3.8%) | |
| Black or African American | 90 (2.5%) | 41 (3.0%) | 49 (2.2%) | |
| Hispanic or Latino | 22 (0.6%) | 12 (0.9%) | 10 (0.4%) | |
| Unknown | 91 (2.5%) | 26 (1.9%) | 65 (2.9%) | |
| **Cancer Category** | | | | |
| Thoracic | 1080 (29.9%) | 397 (29.3%) | 683 (30.3%) | <0.001 |
| Brain/Nervous System | 140 (3.9%) | 46 (3.4%) | 94 (4.2%) | |
| Breast | 122 (3.4%) | 24 (1.8%) | 98 (4.3%) | |
| Gastrointestinal | 411(11.4%) | 149 (11.0%) | 262 (11.6%) | |
| Genitourinary | 387 (10.7%) | 150 (11.1%) | 237 (10.5%) | |
| Gynecologic | 205 (5.7%) | 76 (5.6%) | 129 (5.7%) | |
| Head and Neck | 323 (9.0%) | 191 (14.1%) | 132 (5.9%) | |
| Hematologic | 154 (4.3%) | 82 (6.1%) | 72 (3.2%) | |
| Skin[c] | 750 (20.8%) | 224 (16.5%) | 526 (23.3%) | |
| Others | 35 (1.0%) | 15 (1.1%) | 20 (0.9%) | |
| **Charlson Comorbidity Index[d]** | | | | |
| 0 | 1008 (27.9 %) | 226 (16.7%) | 782 (34.7%) | <0.001 |
| 1-2 | 1542 (42.8%) | 538 (39.7%) | 1004 (44.6%) | |
| 3-4 | 637 (17.7%) | 318 (23.5%) | 319 (14.2%) | |
| >=5 | 420 (11.6%) | 272 (20.1%) | 148 (6.6%) | |
| **ICI Type[e]** | | | | |
| CTLA4 | 66 (1.8%) | 22 (1.6%) | 44 (2.0%) | <0.001 |
| PD-1 | 2731 (75.7%) | 1097 (81.0%) | 1634 (72.5%) | |
| PD-L1 | 439 (12.2%) | 142 (10.5%) | 297 (13.2%) | |
| Combination | 371 (10.3%) | 93 (6.9%) | 278 (12.3%) | |
| **Age at ICI Initiation** | | | | |
| Mean (SD) | 64.2 (13.0) | 65.5 (13.0) | 63.4 (13.0) | <0.001 |
| Median [IQR] | 65.4 [56.7, 73.2] | 67.0 [58.2, 74.4] | 64.4 [55.9, 72.4] | |
| **Year of ICI Initiation** | | | | |
| Earlier than 2016 | 171 (4.7%) | 56 (4.1%) | 115 (5.1%) | 0.241 |
| 2016 | 809 (22.4%) | 314 (23.2%) | 495 (22.0%) | |
| 2017 | 1110 (30.8%) | 394 (29.1%) | 716 (31.8%) | |
| 2018 | 1199 (33.2%) | 465 (34.3%) | 734 (32.6%) | |
| Later than 2018 | 318 (8.8%) | 125 (9.2%) | 193 (8.6%) | |
| **Death Status** | | | | |
| Alive | 1460 (40.5%) | 508 (37.5%) | 952 (42.3%) | 0.006 |
| Death | 2147 (59.5%) | 846 (62.5%) | 1301 (57.7%) | |
| **Duration of Follow-up (days)** | | | | |
| Mean (SD) | 668 (594) | 621 (579) | 696 (602) | <0.001 |
| Median [IQR] | 482 [145, 1088] | 423 [123, 1019] | 535 [161, 1109] | |

[a] Time to cirAE: Time duration from ICI Initiation to cirAE onset in days, only applicable to patients who developed cirAEs.
[b] Patients who developed cirAEs with grades 3 or 4 were grouped together, as only small proportion of patients developed severe cirAEs.

[c] Melanoma comprises 95.1% of this group with the remainder comprised by cutaneous squamous cell carcinoma (4.9%).
[d] ICD codes from visits before ICI initiation were used to calculate Charlson Comorbidity Index.
[e] ICI treatments were separated into four categories: anti-PD-1 (pembrolizumab, nivolumab, cemiplimab), anti-PD-L1 (atezolizumab, avelumab, durvalumab), anti-CTLA4 (ipilimumab), and combination therapy (anti-CTLA4 and either anti-PD-1 or anti-PD-L1).

**eTable4.** Impact of pIDs on cirAE timing.

| | **Non-cutaneous pIDs (N=426)** | **Cutaneous pIDs (N=114)** | **P-value** |
|---|---|---|---|
| **Time from ICI initiation to CirAE (days)** | | | |
| Mean (SD) | 146 (208) | 98 (104) | 0.021 |
| **Logistic Regression Modeling[a]** | | | |
| Beta [95% Confidence Interval] | Reference | -0.11 [-0.21, -0.01] | 0.033 |

[a] The model was adjusted by Race/Ethnicity, Sex, Charlson Comorbidity Index, Age at ICI Initiation, ICI Type, Year of ICI Initiation, and Cancer Category.

**eTable 5.** Associations of pIDs with cirAE development using multivariate Cox Proportional Hazards models, where at least two instances of each pID ICD code were required to be categorized as a pID.

| Pre-existing disease[a] | Hazard Ratio | 95% CI[b] | p-value | q-value[c] | Patients with pIDs[d] | Patients with pIDs and cirAE[e] |
|---|---|---|---|---|---|---|
| Any pID | 1.35 | 1.11, 1.65 | **0.003** | **0.01** | 847 (23.5%) | 166 (19.6%) |
| Cutaneous pIDs | 1.70 | 1.28, 2.26 | **<0.001** | **0.002** | 249 (6.9%) | 60 (24.1%) |
| Non-cutaneous pIDs | 1.21 | 0.96, 1.53 | 0.1 | 0.2 | 598 (15.6%) | 106 (17.7%) |
| Both Cutaneous and Non-cutaneous pIDs | 1.99 | 1.29, 3.08 | **0.002** | **0.011** | 88 (2.4%) | 24 (27.3%) |
| Atopic dermatitis | 5.75 | 2.91, 11.4 | **<0.001** | **<0.001** | 17 (0.5%) | 9 (52.9%) |
| Morphea | 2.58 | 0.63, 10.6 | 0.2 | 0.3 | 9 (0.2%) | 2 (22.2%) |
| Psoriasis | 1.81 | 1.10, 2.97 | **0.019** | **0.045** | 71 (2%) | 17 (23.9%) |
| Vitiligo | 1.02 | 0.14, 7.31 | >0.9 | >0.9 | 5 (0.1%) | 1 (20%) |
| Alopecia areata | 9.01 | 2.18, 37.3 | **0.002** | **0.011** | 2 (0.1%) | 2 (100%) |
| Polymyalgia rheumatica | 2.22 | 1.03, 4.77 | **0.041** | 0.081 | 27 (0.7%) | 7 (25.9%) |
| Systemic sclerosis | 4.87 | 1.52, 15.6 | **0.008** | 0.020 | 6 (0.2%) | 3 (50%) |

[a] Each row corresponds to a separate multivariate Cox Proportional Hazard model to investigate the association of the grouped pre-existing inflammatory diseases (any pID, cutaneous pIDs, non-cutaneous pIDs, and both cutaneous and non-cutaneous pIDs) and individual inflammatory disease with cirAE development. Each model was adjusted by Race/Ethnicity, Sex, Charlson Comorbidity Index, Age at ICI Initiation, ICI Type, Year of ICI Initiation, and Cancer Category. All models have the same reference group, which is the population without any pID diagnosis.

[b] 95% Confidence Interval.

[c] False discovery rate correction for multiple testing.

[d] Patients with pIDs: total number (percentage of the ICI population).

[e] Patients with pIDs and at least one cirAE: total number (percentage of the pID population).

**eTable 6.** Associations of pIDs with cirAE development using multivariate Cox Proportional Hazards models, where only the pID ICD codes within 6 months or 12 months before ICI initiation were included.

| Pre-existing disease[a] | Hazard Ratio | 95% CI[b] | p-value | Patients with pID[c] | Patients with pIDs and CirAE[d] |
|---|---|---|---|---|---|
| **Having diagnoses of pIDs within 12 months before ICI initiation** | | | | | |
| Any pID | 1.18 | 0.97, 1.44 | 0.10 | 883 (28.2%) | 155 (17.6%) |
| Cutaneous pIDs | 1.49 | 1.11, 2.01 | **0.008** | 256 (8.2%) | 55 (21.5%) |
| Non-cutaneous pIDs | 1.05 | 0.83, 1.32 | 0.7 | 627 (20.0%) | 100 (15.9%) |
| Both Cutaneous and Non-cutaneous pIDs | 1.83 | 1.13, 2.98 | **0.014** | 76 (2.4%) | 19 (25.0%) |
| **Having diagnoses of pIDs within 6 months before ICI initiation** | | | | | |
| Any pID | 1.22 | 0.98, 1.50 | 0.069 | 721 (24.2%) | 127 (17.6%) |
| Cutaneous pIDs | 1.62 | 1.16, 2.27 | **0.005** | 178 (6.0%) | 40 (22.5%) |
| Non-cutaneous pIDs | 1.07 | 0.84, 1.37 | 0.6 | 543 (18.3%) | 87 (16.0%) |
| Both Cutaneous and Non-cutaneous pIDs | 1.50 | 0.76, 2.96 | 0.2 | 44 (1.5%) | 9 (20.5%) |

[a] Each row corresponds to a separate multivariate Cox Proportional Hazard model to investigate the association of the grouped pre-existing inflammatory diseases (any pID, cutaneous pIDs, non-cutaneous pIDs, and both cutaneous and non-cutaneous pIDs). Each model was adjusted by Race/Ethnicity, Sex, Charlson Comorbidity Index, Age at ICI Initiation, ICI Type, Year of ICI Initiation, and Cancer Category. All models have the same reference group, which is the population without any pID diagnosis.
[b] 95% Confidence Interval.
[c] Patients with pIDs: total number (percentage of the ICI population).
[d] Patients with pIDs and at least one cirAE: total number (percentage of the pID population).
